# Wearable-measured 24-hour heart rate variability and neovascular age-related macular degeneration: a cross-sectional study

**DOI:** 10.64898/2026.11.20.26368607

**Authors:** Dong-Wouk Park

**Affiliations:** Casey Eye Institute, Oregon Health & Science University, Portland, Oregon, USA

**Author notes:** Corresponding author: Dong-Wouk Park, MD, 3303 S Bond Ave, Building 1, 11th Floor, Portland, OR 97239, USA.

**Keywords:** neovascular age-related macular degeneration, heart rate variability, SDANN, Fitbit, wearable devices, All of Us

## Abstract

**Objective:** To examine wearable heart rate measures in age-related macular degeneration (AMD), reporting an exploratory observation in neovascular AMD.

**Results:** This cross-sectional All of Us analysis used minute-level Fitbit heart rate from adults aged ≥50 years with eye care records. Among the 55 participants with neovascular AMD, the mean date of complete Fitbit recording days fell a median of 1.9 years after the first neovascular AMD diagnosis code. The 7259 comparators had no AMD. The planned primary test, sleeping heart rate with any AMD, was not statistically significant (odds ratio [OR] per 10 beats/min, 1.12; 95% CI, 0.97–1.29). Higher standard deviation of 5-minute average beat intervals (SDANN), a secondary measure, was associated with higher odds of neovascular AMD versus no AMD (OR per 10 ms, 1.12 [95% CI, 1.04–1.21], uncorrected P = 0.002, Bonferroni-adjusted P = 0.051 for 22 comparisons). In an exploratory comparison with non-neovascular AMD, the OR was 1.20 (95% CI, 1.08–1.32). After 24-hour heart rate adjustment, the OR versus no AMD was 1.13 (95% CI, 1.04–1.24). No associations were detected for epiretinal membrane or cataract. This exploratory observation warrants testing in recordings preceding conversion.

## Introduction

Age-related macular degeneration (AMD) is a leading cause of irreversible blindness among older adults in the United States and Western Europe [1,2]. Home monitoring can detect conversion to neovascular AMD, an advanced form of the disease, with less vision lost [3]. Heart rate and heart rate variability, which consumer wearable devices now measure during daily life, are candidate systemic signals, but whether they change before conversion remains uncertain. Adrenergic signaling, which also regulates heart rate, may be relevant. In a mouse model, β-adrenergic blockade reduced the abnormal choroidal vessel growth characteristic of neovascular AMD [4]. Short-term heart rate variability ratios have suggested reduced vagal tone in treated neovascular AMD, although standard time-domain measures did not differ from controls [5]. Resting pulse rate showed no statistically significant association with neovascular AMD in a Scottish cohort [6].

The All of Us Research Program links electronic health records with consumer wearable recordings [7,8]. Minute-level heart rate can be used to estimate the standard deviation of 5-minute average beat intervals (SDANN) over 24 hours. SDANN reflects variation over periods longer than 5 minutes [9], including changes related to activity and the day–night cycle. This cross-sectional analysis examined wearable measures in AMD. Sleeping heart rate with any AMD was the planned primary test, and the neovascular AMD SDANN finding arose in the secondary analysis.

## Methods

The study used the All of Us Registered Tier Dataset v9 (R2025Q4R6). Participants were aged ≥50 years at the mean date of their complete Fitbit days, had an eye diagnosis or procedure code, and had ≥1 complete day of heart rate data. Atrial fibrillation, atrial flutter, and pacemaker codes were exclusions. Analyses remained within the Researcher Workbench. Participant counts of 1–20 and statistics from which those counts could be derived were suppressed in accordance with the All of Us Data and Statistics Dissemination Policy. Code lists and expanded methods appear in Additional file 1.

AMD was identified from diagnosis codes at any date. Neovascular AMD comprised ICD-10-CM H35.32x or ICD-9-CM 362.52, including inactive disease and scar. Any AMD also included non-neovascular and unspecified AMD. Late AMD comprised geographic atrophy or neovascular AMD. Drusen codes alone did not qualify. Participants without AMD codes formed the main comparison group. Non-neovascular AMD, epiretinal membrane, cataract or lens implant, and intravitreal injection without AMD were examined in exploratory comparisons.

A complete day contained heart rate readings of 30–220 beats/min in all 1440 minutes. Each minute’s heart rate was converted to a beat interval in milliseconds by dividing 60 000 by heart rate. Intervals were averaged in 5-minute blocks, and the standard deviation (SD) across blocks gave daily SDANN. Participant values were medians across complete days. This pulse-derived approximation is not an electrocardiographic beat-to-beat measurement. Daily mean heart rate and the SD of 5-minute average heart rates in beats/min were summarized similarly. Sleeping heart rate was the mean during the main sleep period on nights with ≥3 hours of data, summarized across ≥3 such nights.

Logistic regression estimated odds ratios (ORs) per 10 ms of SDANN, per 10 beats/min of sleeping heart rate, and per 1 beat/min of variability computed in beats/min. Models adjusted for age and age squared at the recordings, sex, race and ethnicity, education, income, smoking, body mass index, diabetes, hypertension, heart failure, and heart rate–lowering drugs. Covariates other than age were recorded at enrollment. Medication use covered the preceding year. Categories without cases or comparison participants were excluded from individual models. Table 1 describes participants before these exclusions.

**Table 1.** Characteristics of participants with neovascular AMD and without AMD^a^.

| **Characteristic** | **Neovascular AMD (n = 55)** | **No AMD (n = 7259)** |
|---|---|---|
| Age at the middle of the Fitbit period, median (IQR), y | 76 (71-80) | 67 (59-72) |
| **Sex, %<sup>b</sup>** |  |  |
| Women | 60.0 | 60.6 |
| Men, other, or not recorded | 40.0 | 39.4 |
| **Race and ethnicity, %** | **Not shown<sup>c</sup>** |  |
| Hispanic |  | 6.9 |
| Non-Hispanic White |  | 75.0 |
| Non-Hispanic Black |  | 7.9 |
| Non-Hispanic Asian, other, multiple, or unknown |  | 10.2 |
| **Smoking, %<sup>b</sup>** |  |  |
| Ever (≥100 cigarettes) | 45.5 | 37.7 |
| Never, no answer, or not recorded | 54.5 | 62.3 |
| **Education, %<sup>b</sup>** | **Not shown<sup>c</sup>** |  |
| Less than high school |  | 0.6 |
| High school or equivalent |  | 6.9 |
| Some college |  | 24.5 |
| College graduate or more |  | 58.4 |
| No answer or not recorded |  | 9.5 |
| **Annual household income, %<sup>b</sup>** |  |  |
| ≥\$75 000 | 50.9 | 45.7 |
| <\$75 000, no answer, or not recorded | 49.1 | 54.3 |
| Body mass index at enrollment, median (IQR)<sup>d</sup> | 28.5 (26.8-31.9) | 28.8 (25.1-33.6) |
| Diabetes, %<sup>b</sup> | Not shown<sup>c</sup> | 19.1 |
| Hypertension, %<sup>b</sup> | 52.7 | 48.1 |
| Heart failure, %<sup>b</sup> | Not shown<sup>c</sup> | 3.4 |
| Heart rate–lowering drug, %<sup>b</sup> | Not shown<sup>c</sup> | 16.8 |
| Complete Fitbit days, median (IQR) | 70 (12-149) | 41 (10-122) |
| 24-hour heart rate, median (IQR), beats/min | 69.4 (64.8-75.7) | 72.9 (67.7-78.3) |
| SDANN, median (IQR), ms | 117.5 (97.9-142.0) | 106.1 (86.4-128.2) |
| SD of 5-minute average heart rate, median (IQR), beats/min | 10.3 (8.7-11.5) | 9.9 (8.5-11.4) |
| Daily steps, median (IQR) | 5491 (3044-8052) | 5912 (3982-8388) |
| Sleeping heart rate, median (IQR), beats/min<sup>e</sup> | 60.7 (56.4-67.2) | 64.1 (58.7-69.8) |
| Years from first neovascular AMD code to mean Fitbit recording date, median (IQR) | 1.9 (−0.1 to 4.7) | NA |

Abbreviations: AMD, age-related macular degeneration; IQR, interquartile range; NA, not applicable; SDANN, SD of 5-minute average beat intervals.
<sup>a</sup> All participants in the analysis. Where a category or its complement held 1 to 20 participants, categories were combined or not shown, as program policy requires.
<sup>b</sup> From enrollment records: sex, survey answers, diagnosis codes on or before enrollment, and heart rate–lowering drugs in the year before enrollment. Participants without enrollment physical measurements had none of these recorded; they are counted as not recorded and as without a code or drug, and they were not in the fully adjusted models. Percentages with a code or drug are therefore lower bounds.
<sup>c</sup> Not shown because a category, or its complement, held 1 to 20 participants.
<sup>d</sup> Weight in kilograms divided by height in meters squared, among participants with a measurement.
<sup>e</sup> Participants with at least 3 nights of sleeping heart rate.

The initial analysis comprised 22 distinct comparisons: 7 Fitbit measures (sleeping heart rate, 24-hour heart rate, SDANN, minutes awake, awakenings, sleep efficiency, and total sleep), each in any, late, and neovascular AMD, plus Apple Watch heart rate variability in any AMD; too few participants with late or neovascular AMD had Apple Watch data. All are reported in Additional file 1, Table S2. Sleeping heart rate with any AMD was designated primary before that analysis ran. SDANN by AMD type was secondary. Uncorrected two-sided P values are shown, with the initial findings also assessed by Bonferroni correction for 22 comparisons.

All checks of the neovascular AMD SDANN finding were added after the initial results. They examined mean heart rate, age modeling, physical activity, recorded injections, comorbidity, medication timing, sleep duration, minimum wear time, stricter case definitions, eye care ascertainment, extreme values, and control outcomes. Firth penalized regression addressed sparse-data bias, including a model retaining covariate categories without cases. Some checks had decision rules set before they ran. All were still added after the initial results. Additional file 1 provides the complete estimates, chronology, decision rules, and methods. Analyses were performed in Python within the All of Us Researcher Workbench. No independent validation sample was used.

Analysis code was developed with assistance from Claude (Anthropic) and ChatGPT (OpenAI). The author reviewed and executed the code. Reporting followed the Strengthening the Reporting of Observational Studies in Epidemiology (STROBE) guideline.

## Results

The neovascular AMD comparison included 55 participants with neovascular AMD and 7259 without AMD. Median ages were 76 and 67 years, and median numbers of complete Fitbit days were 70 and 41, respectively (Table 1). Among participants with neovascular AMD, the median interval from the first recorded neovascular AMD code to the mean date of complete Fitbit recording days was 1.9 years (interquartile range, −0.1 to 4.7 years).

Sleeping heart rate, the planned primary test, was not statistically significant for any AMD (OR per 10 beats/min, 1.12; 95% CI, 0.97–1.29; P = 0.11). In the secondary analysis, higher SDANN was associated with higher odds of neovascular AMD (OR per 10 ms, 1.12 [95% CI, 1.04–1.21], uncorrected P = 0.002). The Bonferroni-adjusted P value for 22 comparisons was 0.051, just above 0.05. Across the 21 Fitbit comparisons alone, a grouping defined after the results, the Bonferroni-adjusted P value was 0.049. SDANN was not associated with any AMD (OR, 1.00; 95% CI, 0.96– 1.03). Two other initial comparisons were statistically significant before correction: SDANN with late AMD, which included neovascular AMD (P = 0.009), and minutes awake with any AMD (P = 0.006). None of the 22 comparisons remained significant after correction (Additional file 1, Table S2).

Participants with neovascular AMD had lower median 24-hour heart rate (69.4 versus 72.9 beats/min) and higher median SDANN (117.5 versus 106.1 ms). The exploratory unadjusted OR per 10 ms of SDANN was 1.11 (95% CI, 1.04–1.19, uncorrected P = 0.002). With adjustment for age, age squared, and sex, the OR was 1.09 (95% CI, 1.02–1.17, uncorrected P = 0.02). Both estimates are reported in Additional file 1, Table S3. With 24-hour heart rate adjustment, the SDANN OR was 1.13 (95% CI, 1.04–1.24). Both Firth models gave an OR of 1.12 (95% CI, 1.04–1.21), including the model that retained participants in covariate categories without cases. Variability computed directly in beats/min was associated after full adjustment (OR per 1 beat/min, 1.15; 95% CI, 1.03–1.29), but not without adjustment (OR, 1.07; 95% CI, 0.96–1.18) or with adjustment for age, age squared, and sex (P = 0.06).

Without recorded heart rate–lowering drugs in the year before enrollment, the SDANN OR was 1.10 (95% CI, 1.00–1.22; P = 0.06). Medication analyses timed to the recordings were added after this result; excluding recorded use of any listed drug gave an OR of 1.12 (95% CI, 1.02–1.24). Requiring an eye procedure code in both groups gave an OR of 1.10 (95% CI, 1.01–1.19). Against non-neovascular AMD, the OR was 1.20 (95% CI, 1.08–1.32). SDANN was not associated with epiretinal membrane or cataract/lens implant (Table 2). The full exploratory results are in Additional file 1, Tables S3 and S4.

**Table 2.**
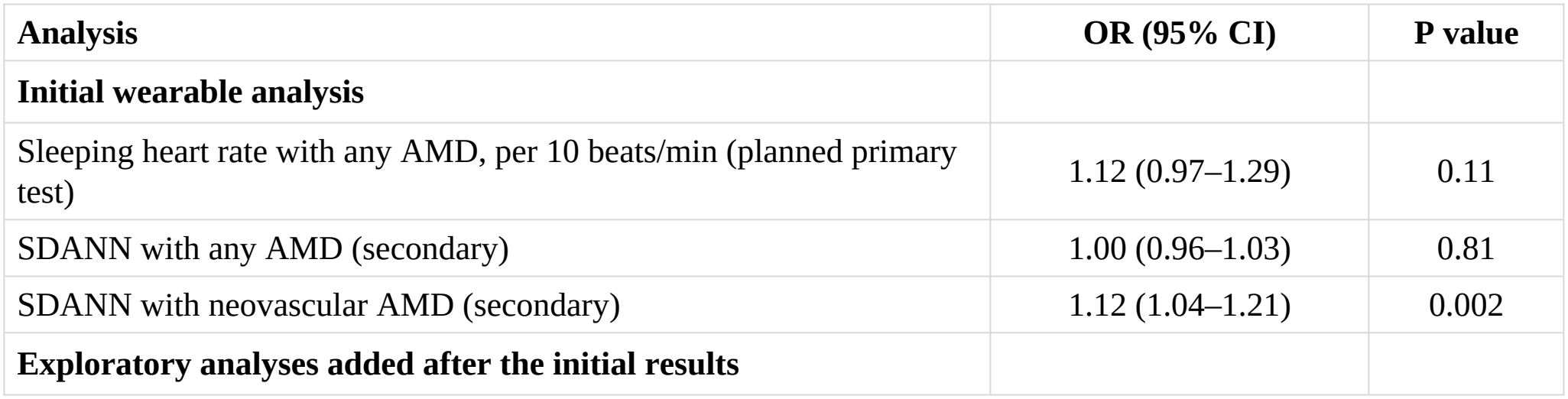
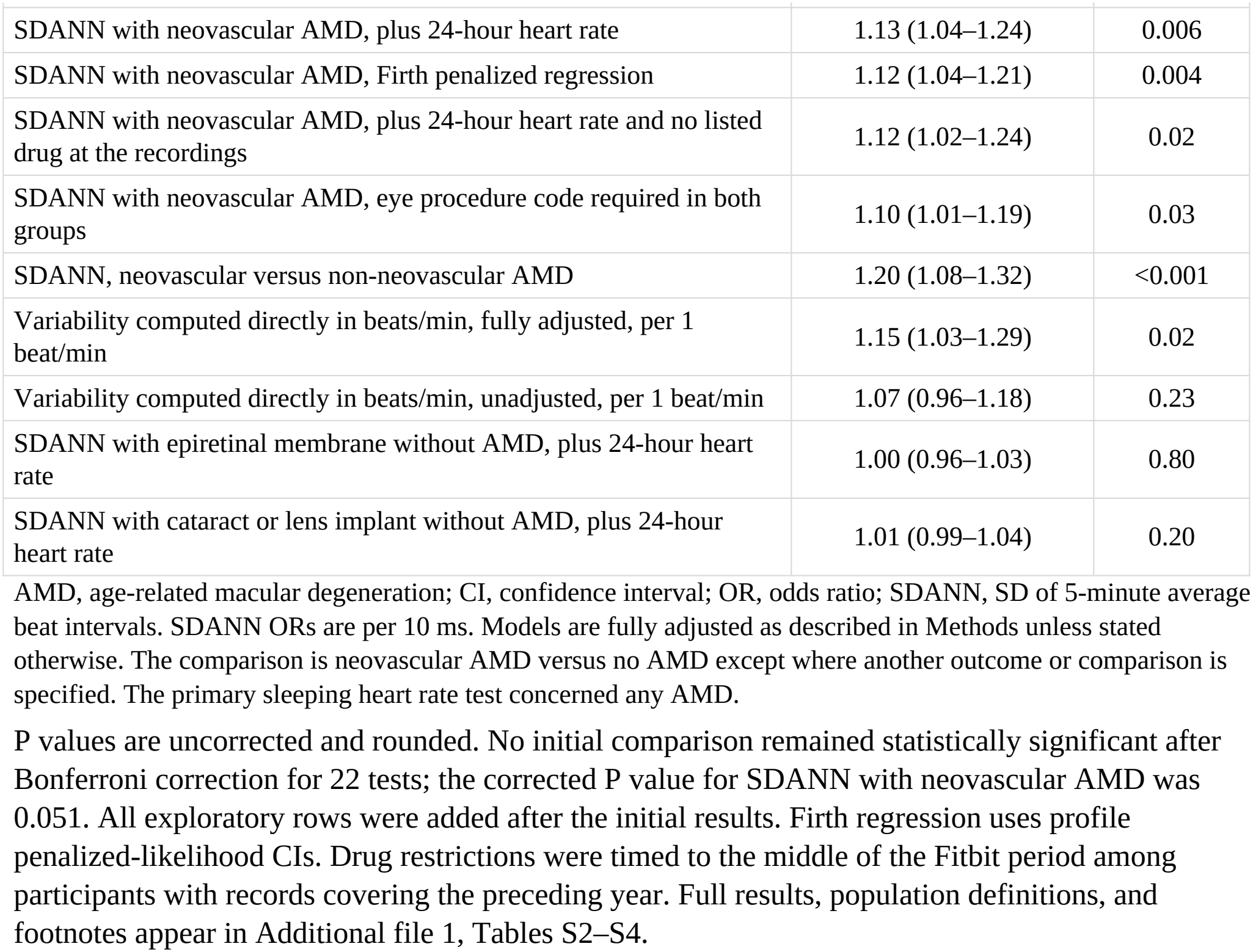
Initial wearable results and selected exploratory analyses.

| **Analysis** | **OR (95% CI)** | **P value** |
|---|---|---|
| **Initial wearable analysis** |  |  |
| Sleeping heart rate with any AMD, per 10 beats/min (planned primary test) | 1.12 (0.97–1.29) | 0.11 |
| SDANN with any AMD (secondary) | 1.00 (0.96–1.03) | 0.81 |
| SDANN with neovascular AMD (secondary) | 1.12 (1.04–1.21) | 0.002 |
| **Exploratory analyses added after the initial results** |  |  |
| SDANN with neovascular AMD, plus 24-hour heart rate | 1.13 (1.04–1.24) | 0.006 |
| SDANN with neovascular AMD, Firth penalized regression | 1.12 (1.04–1.21) | 0.004 |
| SDANN with neovascular AMD, plus 24-hour heart rate and no listed drug at the recordings | 1.12 (1.02–1.24) | 0.02 |
| SDANN with neovascular AMD, eye procedure code required in both groups | 1.10 (1.01–1.19) | 0.03 |
| SDANN, neovascular versus non-neovascular AMD | 1.20 (1.08–1.32) | <0.001 |
| Variability computed directly in beats/min, fully adjusted, per 1 beat/min | 1.15 (1.03–1.29) | 0.02 |
| Variability computed directly in beats/min, unadjusted, per 1 beat/min | 1.07 (0.96–1.18) | 0.23 |
| SDANN with epiretinal membrane without AMD, plus 24-hour heart rate | 1.00 (0.96–1.03) | 0.80 |
| SDANN with cataract or lens implant without AMD, plus 24-hour heart rate | 1.01 (0.99–1.04) | 0.20 |

AMD, age-related macular degeneration; CI, confidence interval; OR, odds ratio; SDANN, SD of 5-minute average beat intervals. SDANN ORs are per 10 ms. Models are fully adjusted as described in Methods unless stated otherwise. The comparison is neovascular AMD versus no AMD except where another outcome or comparison is specified. The primary sleeping heart rate test concerned any AMD.
P values are uncorrected and rounded. No initial comparison remained statistically significant after Bonferroni correction for 22 tests; the corrected P value for SDANN with neovascular AMD was 0.051. All exploratory rows were added after the initial results. Firth regression uses profile penalized-likelihood CIs. Drug restrictions were timed to the middle of the Fitbit period among participants with records covering the preceding year. Full results, population definitions, and footnotes appear in Additional file 1, Tables S2–S4.

## Discussion

Sleeping heart rate with any AMD, the planned primary test, was not statistically significant (OR per 10 beats/min, 1.12; 95% CI, 0.97–1.29); the interval remains compatible with a modest positive association. The secondary analysis identified greater 24-hour beat-interval variability in neovascular AMD. The Bonferroni-adjusted P value was 0.051, just above 0.05. The estimate therefore remains exploratory. Subsequent checks within these data provide no independent confirmation.

This study provides an initial assessment of 24-hour variability derived from wearable heart rate by AMD type in a national cohort. The association against non-neovascular AMD, together with the absence of associations for epiretinal membrane and cataract, makes a uniform eye-care ascertainment or device artifact less consistent with the observed pattern. Differences in treatment, sensor performance, or wear patterns across diagnoses could still have influenced the results. The current release included too few eligible participants with Fitbit recordings before their first neovascular AMD code to test whether greater 24-hour variability precedes documented diagnosis.

At a lower mean heart rate, the same fluctuation in beats/min produces a larger fluctuation in beat intervals. Using the group medians and assuming equal variability in beats/min, the difference in heart rate would correspond to about 10% higher SDANN in neovascular AMD, close to the observed 11% difference in median SDANN. The association persisted after adjustment for mean heart rate (OR, 1.13; 95% CI, 1.04–1.24). SDANN summarizes how average beat intervals vary across 5-minute blocks over 24 hours. Averaging within each block smooths rapid beat-to-beat fluctuations, so SDANN primarily reflects slower changes across the day [9]. A higher SDANN value therefore offers limited information about short-term vagal modulation. The SDANN association persisted after adjustment for daily steps. In a separate model, the association with approximated variability in beats/min persisted after adjustment for the day–night difference in heart rate (Additional file 1, Table S3). Because the direct beats/min measure was associated only after full adjustment, the contributions of heart rate scaling and covariates remain unresolved.

Sleep apnea has been associated with neovascular AMD in a small cross-sectional study [10], and a small study found that intravitreal bevacizumab blunted nighttime blood pressure dipping [11].

Clinical β-blocker findings conflict [12–15]. Recorded comorbidity and medication checks changed the estimate little, but cannot exclude effects of treatment, unrecorded drugs, visual disability, or daily routines. These results do not support wearable screening or prediction of neovascular AMD. Testing recordings obtained before conversion would be needed to establish temporal ordering.

### Limitations

For most participants with neovascular AMD, the mean Fitbit recording date followed the first recorded neovascular AMD code. The first code can postdate disease onset. Earlier treatment, injections outside participating systems, and changes caused by visual disability remain possible explanations. Visual acuity was unavailable. Laterality and disease activity were not analyzed. Non-neovascular AMD differs in treatment, visit frequency, and vision, limiting interpretation of that comparison.

Only 55 participants had neovascular AMD, with fewer than three cases per estimated coefficient in the initial model. The similar Firth estimates still leave uncertainty from sparse data and selection due to missing covariates. Selecting a statistically significant result from 22 comparisons can inflate its magnitude. Model-specific case counts were suppressed to prevent disclosure of small cells.

Diagnosis codes were not validated against retinal images, and undetected AMD in comparison participants could cause misclassification.

Minute-level optical heart rate has not been validated here against electrocardiographic SDANN. Undiagnosed arrhythmias, ectopic beats, and device differences could affect measurements.

Covariates were recorded at enrollment rather than contemporaneously, and medication records may miss β-blocker use. Requiring complete 24-hour recordings selects for continuous wear. Participants with neovascular AMD contributed a median of 70 complete Fitbit days versus 41 among those without AMD. Requiring at least 3 or 7 complete days changed the estimate little (Additional file 1, Table S3), but differences in activity or wear could still bias the association.

Residual confounding remains possible. For a 1-SD increase in SDANN (33.2 ms), the OR was 1.47 (95% CI, 1.15–1.89), as reported in Additional file 1. The E-values were 2.30 for that estimate and 1.57 for the lower confidence limit [16]. These sensitivity measures do not address reverse causation, selection, or measurement error. The many checks added after the initial results were neither prespecified confirmatory tests nor external replication. The finding remains hypothesis generating and requires evaluation in recordings preceding neovascular AMD.

## Data Availability

This study used data from the All of Us Research Program's Registered Tier Dataset v9 (R2025Q4R6), available to authorized users on the Researcher Workbench. Participant-level data cannot be redistributed by the author. Reportable aggregate results and code lists are included in this article and Additional file 1.

## List of abbreviations

AMD: age-related macular degeneration
CI: confidence interval
ICD-9-CM and ICD-10-CM: International Classification of Diseases, Ninth and Tenth Revisions, Clinical Modification
OR: odds ratio
SD: standard deviation
SDANN: standard deviation of 5-minute average beat intervals
STROBE: Strengthening the Reporting of Observational Studies in Epidemiology.

## Declarations

### Ethics approval and consent to participate

All of Us participants provided written informed consent, signed electronically, under the All of Us institutional review board. The Oregon Health & Science University Institutional Review Board determined that this secondary analysis of deidentified Registered Tier data was not research involving human subjects and that IRB review and approval were not required (STUDY00030932).

### Consent for publication

Not applicable. No identifiable individual participant information is reported.

### Authors’ contributions

DWP conceived and designed the study, performed the analyses, interpreted the findings, and drafted and revised the manuscript. The author read and approved the final manuscript.

## Acknowledgements

We gratefully acknowledge All of Us participants for their contributions, without whom this research would not have been possible. We also thank the National Institutes of Health’s All of Us Research Program for making available the participant data examined in this study.

## Use of artificial intelligence

Claude (Anthropic) and ChatGPT (OpenAI) were used to assist in developing and checking the initial and exploratory analysis code, structuring sensitivity checks, conducting bibliographic searches, and editing the manuscript and supplementary materials for clarity, style, and grammar. The author executed and reviewed all analyses within the All of Us Researcher Workbench and takes full responsibility for the integrity of the data, analyses, references, and final manuscript.

## Additional file

Additional file 1. Expanded methods and supplementary tables. Format: DOCX. Contains the detailed wearable and exploratory analysis methods, Table S1 code lists, Table S2 all 22 initial comparisons and one restricted analysis, Table S3 the complete association and sensitivity results, and Table S4 the decision rules and their readings.

