## Supplements for "Wearable-measured 24-hour heart rate variability and neovascular age-related macular degeneration: a cross-sectional study"

This file provides the code definitions, full initial comparisons, exploratory estimates, and decision rules supporting the research note. References in brackets refer to the main manuscript reference list.

### Expanded methods

#### Data source and participants

The All of Us Research Program enrolls adults across the United States and links electronic health records, surveys, physical measurements and, for participants who share them, Fitbit data [7,8]. This cross-sectional study used the Registered Tier Dataset v9 (R2025Q4R6) on the Researcher Workbench. Analyses were run within the Workbench, and no participant-level data left it. Counts from 1 to 20, and statistics from which such counts could be derived, are not reported, as program policy requires.

Participants were included if they were aged 50 years or older at the middle of their Fitbit period, had eye care in their records, and had at least 1 complete day of Fitbit heart rate data. Eye care was defined as an eye diagnosis code (International Classification of Diseases, Tenth Revision, Clinical Modification [ICD-10-CM] H00-H59 or Ninth Revision [ICD-9-CM] 360-379) or a Current Procedural Terminology (CPT) code for an eye examination, ophthalmic imaging, visual field testing, or intravitreal injection. The eye-care requirement was intended to improve ascertainment of AMD. Participants with an atrial fibrillation or flutter code (ICD-10-CM I48; ICD-9-CM 427.31 or 427.32) or a pacemaker code (ICD-10-CM Z95.0; ICD-9-CM V45.01) were excluded, because these conditions alter heart rate variability.

AMD was identified from diagnosis codes at any date. Any AMD was defined by ICD-10-CM codes H35.30, H35.31x, or H35.32x or ICD-9-CM codes 362.50 to 362.52, and neovascular AMD by H35.32x (any stage, including inactive disease and scar) or 362.52. The comparison group had no AMD code; drusen codes alone did not count as AMD. Two control outcomes, epiretinal membrane and cataract or lens implant, were each defined in participants without AMD and compared with participants without AMD who lacked the condition being tested. Codes are listed in Table S1.

#### Wearable measures and covariates

Fitbit devices estimate heart rate optically, and All of Us provides it at 1-minute resolution. A complete day had heart rate data (readings of 30 to 220 beats/min) in all 1440 minutes. On each complete day, each minute's heart rate was converted to a beat interval (60 000/heart rate), the intervals were averaged in 5-minute blocks, and the SD of the block averages gave that day's SDANN [9]. This approximates electrocardiographic SDANN from pulse rate. The day's 24-hour heart rate was the mean of the 5-minute average heart rates. Each participant's values were summarized as the median across complete days, and the middle of the Fitbit period was the mean date of those days. Variability in beats per minute was the SD of the 5-minute average heart rates, computed on the same complete days and summarized the same way. It was first approximated as

SDANN × heart rate²/60 000; because the approximation correlated with the directly computed value at only r = 0.95, the direct value, computed after the initial results, is the one reported in the text; check 10 used only the approximation. Sleeping heart rate was the mean heart rate during the main sleep period on nights with at least 3 hours of data, summarized as the median across nights for participants with at least 3 such nights.

Models adjusted for age and its square at the middle of the Fitbit period, sex, race and ethnicity, education, income, smoking, body mass index, diabetes, hypertension, heart failure, and heart rate–lowering drug use in the year before enrollment (oral or ophthalmic β-blockers, diltiazem, verapamil, digoxin, amiodarone, or clonidine). Covariates other than age were taken as at enrollment (Table 1). Participants in a covariate category with no cases or no comparison participants were excluded from that model, because that category's coefficient cannot be estimated. In the initial neovascular AMD model, these categories were other sex, ever smoking with unknown current status, missing smoking status, and other, multiple, or unknown race and ethnicity.

**Initial analysis and exploratory checks**

Logistic regression estimated odds ratios (ORs) per 10 ms of SDANN, per 1 beat/min of variability in beats per minute, and per 10 beats/min of sleeping heart rate. The wearable analysis named sleeping heart rate and any AMD as its primary test before it was run; SDANN by AMD type was secondary, and Table S2 shows all 22 distinct comparisons. Clinic heart rate and blood pressure before neovascular AMD are outside the scope of this report.

Checks of the SDANN association with neovascular AMD were added after the initial results; all but check 8 had a decision rule set before it ran (Table S4). For example, an alternative explanation was accepted if it moved the OR at least halfway to 1 on the log scale. The checks (1) added 24-hour heart rate to the model; (2) modeled variability in beats per minute; (3) replaced age and its square with 5-year age bands; (4) restricted the analysis to participants aged 65 years or older; (5) added median daily steps, from days with 100 or more steps; (6) excluded participants with a recorded intravitreal injection (CPT 67028 or an anti-VEGF drug code) within 1 year of the middle of their Fitbit period; (7) added sleep apnea and 6 other cardiovascular and renal diagnoses ever coded; (8) restricted the analysis to participants taking no heart rate–lowering drug in the year before enrollment, with 24-hour heart rate in the model; (9) added 24-hour heart rate and nightly hours asleep and awake in bed; and (10) added the day-night difference in heart rate (awake minus sleeping heart rate) to the model of approximated variability in beats per minute (populations in Table S3).

Prompted by the result of check 8, drug use was then assessed at the time of the recordings, with 24-hour heart rate in the model. A drug was considered in use if a prescription period (fills 180 days or less apart) began on or before the middle of the Fitbit period and included a fill in the preceding year; this could be determined only for participants whose records covered that year. Three restrictions were fitted: no listed drug, no listed oral drug (with ophthalmic β-blockers as a covariate), and no β-blocker (with the other 5 drugs as a covariate).

Four further checks were added later, each also with a rule set before it ran. Because the neovascular AMD model had fewer than 3 cases per estimated coefficient, it was refitted with Firth penalized logistic regression to reduce small-sample bias, with profile penalized-likelihood CIs. One fit used the same participants as the initial model. A second fit retained participants in covariate categories without cases. Both fits required complete model covariates. Participants with neovascular AMD were compared with those with an AMD code but no neovascular AMD code (non-neovascular AMD). The analysis was restricted to participants with at least 3, and at least 7, complete days. Finally, both groups were restricted to participants with an eye examination, imaging, visual field, or intravitreal injection code, so that no participant qualified for eye care through a diagnosis code alone.

A final set of checks was added after all other results had been seen (Table S4). Four had rules set before they ran: SDANN winsorized at the 1st and 99th percentiles; cases redefined by codes on 2 or more dates or by a code plus an injection code; an injection code in participants without AMD, as a third control outcome; and SDANN against years since the first code, among cases. Unadjusted and age-and-sex models, non-neovascular AMD vs no AMD, the OR per SD of SDANN in participants without AMD, and apparent c-statistics, not corrected for optimism, were described.

In a check added after the initial results, SDANN was also related to the first 2 control outcomes, with 24-hour heart rate in the model. Analyses used Python 3.12.14 with pandas 2.3.3, NumPy 2.5.3, statsmodels 0.15.0, and SciPy 1.18.1 (Firth regression was coded for this study). Analysis code was developed with assistance from Claude (Anthropic) and ChatGPT (OpenAI). The author reviewed and executed the code. Two-sided $P < 0.05$ was considered statistically significant. P values were not corrected for multiple comparisons, except that the main result is also given after Bonferroni correction for the 22 distinct comparisons. The study followed the Strengthening the Reporting of Observational Studies in Epidemiology (STROBE) reporting guideline.

#### Additional exploratory results

Among participants without AMD, sleep apnea was associated with lower SDANN: the age- and sex-adjusted difference was −7.75 ms (95% CI, −9.38 to −6.12), and the difference with additional heart rate adjustment was −6.59 ms (95% CI, −7.93 to −5.26). Among neovascular AMD cases, SDANN showed no trend with years since the first code (−0.63 ms per year; 95% CI, −2.15 to 0.89).

The OR per SD of SDANN in participants without AMD (33.2 ms) was 1.47 (95% CI, 1.15–1.89). Apparent c-statistics were 0.61 for SDANN alone, 0.78 for the covariates, and 0.80 with SDANN added. These were not corrected for optimism or externally validated. E-values were 2.30 for the point estimate and 1.57 for its lower confidence limit, using the OR as an approximation to a risk ratio because neovascular AMD was rare [16].

#### Table S1 Code lists

| Definition | ICD-10-CM | ICD-9-CM | Procedure or drug codes |
|---|---|---|---|
| Any AMD | H35.30, H35.31x, H35.32x | 362.50, 362.51, 362.52 | — |
| Neovascular AMD | H35.32x, any laterality and stage (active, inactive, or scar) | 362.52 | — |

| Definition | ICD-10-CM | ICD-9-CM | Procedure or drug codes |
|---|---|---|---|
| Non-neovascular AMD (AMD without a neovascular code) | H35.30 or H35.31x, and no H35.32x | 362.50 or 362.51, and no 362.52 | — |
| Late AMD (Table S2): geographic atrophy or neovascular AMD | H35.31x3 or H35.31x4 (geographic atrophy), or H35.32x | 362.52 (no ICD-9-CM code was used for geographic atrophy) | — |
| Drusen (not counted as AMD) | H35.36x | 362.57 | — |
| Epiretinal membrane (control outcome) | H35.37x | 362.56 | — |
| Cataract or lens implant (control outcome) | H25.x; H26.x except H26.4x; H27.0x; Z96.1; Z98.4x | 366.x except 366.5x; 379.31; V43.1; V45.61 | — |
| Eye care: eye diagnosis | H00-H59 | 360-379 | — |
| Eye care: eye procedure (also the eye procedure check) | — | — | CPT 92002, 92004, 92012, 92014 (examination); 92132, 92133, 92134, 92137 (optical coherence tomography); 92250 (fundus photography); 92235, 92240, 92242 (angiography); 92201, 92202 (extended ophthalmoscopy); 92081, 92082, 92083 (visual field); 67028 (intravitreal injection) |
| Intravitreal injection (check 6) | — | — | CPT 67028; HCPCS J0178, J0177 (aflibercept), J2778 (ranibizumab), Q5124, Q5128 (ranibizumab biosimilars), J2777 (faricimab), J0179 (brolucizumab); bevacizumab injections are captured through CPT 67028 (its drug codes, J9035 and C9257, were not used) |
| Atrial fibrillation or flutter (exclusion) | I48.x | 427.31, 427.32 | — |
| Pacemaker (exclusion) | Z95.0 | V45.01 | — |
| Diabetes (covariate) | E08-E13 | 250.x | — |
| Hypertension (covariate) | I10-I16 | 401-405 | — |
| Heart failure (covariate; check 7) | I50.x | 428.x | — |
| Heart rate–lowering drugs (covariate) | — | — | ATC C07 (β-blockers) and S01ED (ophthalmic β-blockers); RxNorm ingredients diltiazem, verapamil, digoxin, amiodarone, and clonidine |
| Sleep apnea (check 7) | G47.33 | 327.23 | — |
| Coronary disease (check 7) | I20-I25 | 410-414 | — |

| Definition | ICD-10-CM | ICD-9-CM | Procedure or drug codes |
|---|---|---|---|
| Cerebrovascular disease (check 7) | I60-I69 | 430-438 | — |
| Peripheral artery disease (check 7) | I70.2x, I73.9x | 440.2x, 443.9 | — |
| Chronic kidney disease (check 7) | N18.x | 585.x | Or estimated glomerular filtration rate below 60 mL/min/1.73 m² from the last creatinine in the 3 years up to enrollment |
| Low blood pressure (check 7) | I95.x | 458.x | — |

Abbreviations: AMD, age-related macular degeneration; ATC, Anatomical Therapeutic Chemical classification; CPT, Current Procedural Terminology; HCPCS, Healthcare Common Procedure Coding System; ICD-9-CM and ICD-10-CM, International Classification of Diseases, Ninth and Tenth Revisions, Clinical Modification.

An x stands for any further characters. Diagnosis codes came from condition and observation records, by source code. Participants’ medians across complete days were computed with an approximate quantile function; recomputing them changed SDANN by less than 1 ms for 99% of participants.

#### Table S2 All 22 initial wearable comparisons and one restricted analysis

| Measure | Group vs no AMD | Estimate (95% CI) | P value |
|---|---|---|---|
| Sleeping heart rate, per 10 beats/min (planned primary test) | Any AMD | 1.12 (0.97-1.29) | 0.11 |
| | Late AMD (geographic atrophy or neovascular) | 0.76 (0.52-1.10) | 0.15 |
| | Neovascular AMD | 0.72 (0.48-1.07) | 0.11 |
| 24-hour heart rate, per 10 beats/min | Any AMD | 1.07 (0.92-1.23) | 0.39 |
| | Late AMD | 0.80 (0.55-1.17) | 0.25 |
| | Neovascular AMD | 0.76 (0.51-1.15) | 0.20 |
| SDANN, per 10 ms | Any AMD | 1.00 (0.96-1.03) | 0.81 |
| | Late AMD | 1.10 (1.03-1.19) | 0.009 |
| | Neovascular AMD | 1.12 (1.04-1.21) | 0.002 |
| Minutes awake per night, per 30 min | Any AMD | 0.75 (0.61-0.92) | 0.006 |
| | Late AMD | 0.64 (0.39-1.05) | 0.08 |
| | Neovascular AMD | 0.73 (0.43-1.24) | 0.24 |
| Awakenings per night, per 5 | Any AMD | 0.81 (0.56-1.17) | 0.27 |
| | Late AMD | 0.99 (0.40-2.46) | 0.98 |
| | Neovascular AMD | 1.17 (0.44-3.07) | 0.76 |
| Sleep efficiency, per 5 percentage points | Any AMD | 1.04 (0.87-1.23) | 0.70 |

| Measure | Group vs no AMD | Estimate (95% CI) | P value |
|---|---|---|---|
| | Late AMD | 0.90 (0.69-1.17) | 0.42 |
| | Neovascular AMD | 0.88 (0.67-1.15) | 0.35 |
| Total sleep, per hour | Any AMD | 1.05 (0.95-1.15) | 0.35 |
| | Late AMD | 1.09 (0.86-1.38) | 0.47 |
| | Neovascular AMD | 1.13 (0.88-1.45) | 0.35 |
| Apple Watch SDNN, ratio of geometric means (AMD over no AMD) | Any AMD | 0.95 (0.87-1.03) | 0.21 |
| Restricted analysis: sleeping heart rate, no heart rate–lowering drug | Any AMD | 1.16 (0.99-1.36) | 0.07 |

Abbreviations: AMD, age-related macular degeneration; OR, odds ratio; SDANN, SD of 5-minute average beat intervals; SDNN, SD of normal beat intervals.

Models used the covariates listed in Methods, with participants without AMD as the reference group. The restricted sleeping heart rate model omitted the drug-use covariate. Late AMD: a geographic atrophy or neovascular AMD code. Minutes awake, awakenings, sleep efficiency and total sleep from nights with 3 to 16 hours of main sleep, in participants with 7 or more such nights. Age for these measures was assessed at the mean date of eligible sleep recordings. Sleeping heart rate was defined as in Methods. The Apple Watch comparison used a separate eligible sample, with age assessed at the mean recording date. Late and neovascular AMD were not compared for the Apple Watch because too few participants with those codes had Apple Watch data. The estimate is a ratio of geometric means from a linear model of log SDNN. Case counts are not shown, as program policy requires. Table entries report uncorrected P values. None of the 22 initial comparisons remained statistically significant after Bonferroni correction. For SDANN with neovascular AMD, Bonferroni-corrected P = 0.051 (0.0511 in the recorded analysis output).

#### Table S3 Odds ratios per 10 ms of SDANN unless otherwise stated[a]

| Analysis | OR (95% CI) | P value |
|---|---|---|
| **Initial analysis** | | |
| Neovascular AMD vs no AMD | 1.12 (1.04-1.21) | 0.002 |
| Neovascular AMD vs no AMD, adjusted for age, age squared, and sex only | 1.09 (1.02-1.17) | 0.02 |
| Any AMD vs no AMD | 1.00 (0.96-1.03) | 0.81 |
| **Checks added after the initial results[b]** | | |
| Plus 24-hour heart rate | 1.13 (1.04-1.24) | 0.006 |
| Plus 24-hour heart rate; no heart rate–lowering drug in the year before enrollment[c] | 1.10 (1.00-1.22) | 0.06 |
| Age in 5-year bands | 1.12 (1.04-1.21) | 0.003 |

| Analysis | OR (95% CI) | P value |
|---|---|---|
| Age 65 years or older only | 1.12 (1.04-1.22) | 0.004 |
| Plus daily steps | 1.13 (1.05-1.23) | 0.002 |
| No recorded intravitreal injection within 1 year of the recordings | 1.12 (1.03-1.23) | 0.01 |
| Plus sleep apnea and 6 other diagnoses[d] | 1.12 (1.04-1.20) | 0.004 |
| Plus 24-hour heart rate; no listed drug at the recordings[e] | 1.12 (1.02-1.24) | 0.02 |
| Plus 24-hour heart rate; no listed oral drug at the recordings[e] | 1.14 (1.03-1.25) | 0.01 |
| Plus 24-hour heart rate; no β-blocker at the recordings[e] | 1.13 (1.02-1.24) | 0.02 |
| Plus 24-hour heart rate and nightly hours asleep and awake in bed[f] | 1.13 (1.04-1.24) | 0.007 |
| Firth penalized regression[g] | 1.12 (1.04-1.21) | 0.004 |
| Firth penalized regression, retaining categories without cases[g] | 1.12 (1.04-1.21) | 0.004 |
| 3 or more complete days | 1.11 (1.03-1.21) | 0.007 |
| 7 or more complete days | 1.11 (1.02-1.21) | 0.01 |
| Eye procedure code required in both groups[h] | 1.10 (1.01-1.19) | 0.03 |
| Neovascular AMD vs non-neovascular AMD[i] | 1.20 (1.08-1.32) | <0.001 |
| Neovascular AMD vs non-neovascular AMD, plus 24-hour heart rate[c,i] | 1.22 (1.07-1.38) | 0.002 |
| **Checks: variability in beats per minute, OR per 1 beat/min[j]** | | |
| Computed directly | 1.15 (1.03-1.29) | 0.02 |
| Computed directly, plus 24-hour heart rate[c] | 1.16 (1.03-1.29) | 0.01 |
| Approximated | 1.18 (1.05-1.31) | 0.004 |
| Approximated, plus the day-night difference in heart rate[k] | 1.25 (1.04-1.49) | 0.01 |
| **Control outcomes, added after the initial results, plus 24-hour heart rate[l]** | | |
| Epiretinal membrane without AMD | 1.00 (0.96-1.03) | 0.80 |
| Cataract or lens implant without AMD | 1.01 (0.99-1.04) | 0.20 |
| Intravitreal injection code without AMD[m] | 0.89 (0.78-1.02) | 0.10 |
| **Final checks, added after all other results[m]** | | |
| Neovascular AMD vs no AMD, unadjusted[c] | 1.11 (1.04-1.19) | 0.002 |
| SDANN winsorized at the 1st and 99th percentiles | 1.14 (1.05-1.24) | 0.002 |
| Cases with neovascular AMD codes on 2 or more dates | 1.11 (1.02-1.21) | 0.02 |
| Cases with a neovascular AMD code and an intravitreal injection code | 1.12 (1.02-1.24) | 0.02 |
| Non-neovascular AMD vs no AMD[c] | 0.97 (0.94-1.01) | 0.17 |
| Non-neovascular AMD vs no AMD, plus 24-hour heart rate[c] | 0.98 (0.94-1.03) | 0.47 |

| Analysis | OR (95% CI) | P value |
| --- | --- | --- |
| Variability in beats per minute (direct), per 1 beat/min, unadjusted[c] | 1.07 (0.96-1.18) | 0.23 |
| Variability in beats per minute (direct), per 1 beat/min, adjusted for age, age squared, and sex only[c] | 1.11 (1.00-1.23) | 0.06 |
| Variability in beats per minute (direct), per 1 beat/min, winsorized[c] | 1.16 (1.02-1.32) | 0.03 |

Abbreviations: AMD, age-related macular degeneration; OR, odds ratio; SDANN, SD of 5-minute average beat intervals.

[a] Participants aged 50 years or older at the middle of their Fitbit period, with eye care; the comparison group had no AMD code (55 with neovascular AMD; 7259 without AMD, of whom 6551 had complete covariates and 5950 entered the initial neovascular AMD model). Logistic regression adjusted for the covariates listed in Methods unless stated otherwise.

[b] Each with a decision rule set before it ran, except where noted (rules and readings in Table S4); for example, an explanation required the OR to move at least halfway to 1 on the log scale.

[c] No decision rule was set before this check ran.

[d] Sleep apnea, coronary disease, cerebrovascular disease, peripheral artery disease, chronic kidney disease, low blood pressure, and heart failure, ever coded.

[e] Prompted by the P = 0.06 result. Drug use at the middle of the Fitbit period, in participants whose records covered the year before it (reference estimate in these participants, with drug use at enrollment: 1.13; 95% CI, 1.03-1.24). Oral-drug row: ophthalmic β-blockers as a covariate. β-Blocker row: the other 5 drugs as a covariate.

[f] Participants with 7 or more nights of sleep data (reference estimate, with 24-hour heart rate, 1.14; 95% CI, 1.04-1.25).

[g] Penalized logistic regression with profile penalized-likelihood CIs. Both models required complete model covariates. The first used the same participants as the initial model. The second also retained participants in covariate categories without cases.

[h] An eye examination, imaging, visual field, or intravitreal injection code at any date.

[i] Comparison group: participants with an AMD code but no neovascular AMD code.

[j] SD of the 5-minute average heart rates, computed directly (after the initial results) or approximated as SDANN × heart rate$^2$/60 000 (the earlier checks).

[k] Participants with sleep, waking, and 24-hour summaries (reference estimate, approximated variability alone, 1.17; 95% CI, 1.05-1.32).

[l] Each analysis was restricted to participants without AMD. The comparison group lacked the condition or procedure being tested.

[m] Added after all other results had been seen, each with a decision rule set before it ran unless marked [c] (rules and readings in Table S4).

### Table S4 Decision rules for checks added after the initial results

Every check below was added after the initial wearable results were known. Setting a rule before an individual check ran does not make that check part of the original analysis plan. Labels such as “robust,” “not explained,” and “specific” reproduce the rule-based readings; they do not establish causality, eliminate confounding, or demonstrate disease specificity.

| Check | Rule set before the check ran | Reading |
|---|---|---|
| Plus 24-hour heart rate (1), and variability in beats per minute, approximated (2) | More than heart rate level if the OR stays above 1 with a 95% CI excluding 1 in both | Met: 1.13 (1.04-1.24); 1.18 (1.05-1.31) |
| Variability in beats per minute, computed directly (2; added later) | Holds if the OR is above 1 with a 95% CI excluding 1 | Met: 1.15 (1.03-1.29) |
| 5-year age bands (3), and age 65 years or older (4) | Robust if the OR stays above 1 with a 95% CI excluding 1 in both | Met: 1.12 (1.04-1.21); 1.12 (1.04-1.22) |
| Plus daily steps (5) | Explained by activity if the OR moves at least halfway to 1 | Not explained: 1.13 (1.05-1.23) |
| No recorded intravitreal injection within 1 year (6) | Explained by treatment if the association appears only in participants with an injection | Not explained: 1.12 (1.03-1.23) without an injection |
| Plus sleep apnea, and plus all 7 diagnoses (7) | Explained if either addition moves the OR at least halfway to 1; robust if, with all 7, the OR stays above 1 with a 95% CI excluding 1 | Not explained; robust: 1.12 (1.04-1.21); 1.12 (1.04-1.20) |
| No heart rate–lowering drug in the year before enrollment (8) | None set | Described: 1.10 (1.00-1.22) |
| Drug use at the recordings: no listed drug, no listed oral drug, no β-blocker | Explained by heart rate–lowering drugs if any restriction moves the OR at least halfway to 1 from the reference in the same participants (1.13) | Not explained: 1.12; 1.14; 1.13 |
| Plus nightly hours asleep and awake in bed (9) | Explained by sleep length if the OR moves at least halfway to 1 from the reference in the same participants (1.14) | Not explained: 1.13 (1.04-1.24) |
| Plus the day-night difference in heart rate, approximated variability (10) | Carried by the day-night difference if the OR moves at least halfway to 1 from the reference in the same participants (1.17) | Not carried: 1.25 (1.04-1.49) |
| Firth penalized regression | Sparse-data bias is material if the OR moves at least halfway to 1. The result is insensitive to these exclusions if retaining covariate categories without cases gives an OR above 1 with a 95% CI excluding 1. | Neither estimate moved halfway to 1. The OR was 1.12 (1.04-1.21) in both models. |
| Against non-neovascular AMD | Specific to neovascular AMD within AMD if the OR is above 1 with a 95% CI excluding 1 | Met: 1.20 (1.08-1.32) |
| At least 3, and at least 7, complete days | Robust to wear time if both ORs are above 1 with 95% CIs excluding 1 | Met: 1.11 (1.03-1.21); 1.11 (1.02-1.21) |
| Eye procedure code in both groups | Explained by eye care ascertainment if the OR moves at least halfway to 1 | Not explained: 1.10 (1.01-1.19) |

| Check | Rule set before the check ran | Reading |
|---|---|---|
| Control outcomes: epiretinal membrane; cataract or lens implant | A general pattern if either OR is at or above the halfway mark (from 1.13, with 24-hour heart rate) with a 95% CI excluding 1; specific to neovascular AMD if both are below it | Specific: 1.00 (0.96-1.03); 1.01 (0.99-1.04) |
| SDANN winsorized at the 1st and 99th percentiles | Explained by extreme values if the OR is at or below the halfway mark (1.06) | Not explained: 1.14 (1.05-1.24) |
| Cases with neovascular AMD codes on 2 or more dates; cases with a code and an intravitreal injection code | Explained by a loose case definition if either OR is at or below the halfway mark (1.06) | Not explained: 1.11 (1.02-1.21); 1.12 (1.02-1.24) |
| Control outcome: intravitreal injection code without AMD, plus 24-hour heart rate | Treatment supported as an explanation if the OR is at or above the halfway mark of the neovascular estimate in the same model (1.06, from 1.13) with a 95% CI excluding 1; not supported if the upper 95% limit is below that mark | Not supported: 0.89 (0.78-1.02) |
| Years since the first neovascular AMD code, cases only (linear regression adjusted for age, age squared, and 24-hour heart rate) | A consequence of the disease is suggested if SDANN rises with years since the first code (95% CI excluding 0) | No trend: −0.63 ms per year (−2.15 to 0.89) |
| Unadjusted models (SDANN; beats per minute); age-and-sex model and winsorizing for beats per minute; non-neovascular AMD vs no AMD; OR per SD; c-statistics; sleep apnea difference also adjusted for heart rate; 24-hour heart rate added to the direct measure, to neovascular vs non-neovascular AMD, and to non-neovascular AMD vs no AMD | None set (described) | 1.11 (1.04-1.19); 1.07 (0.96-1.18); 1.11 (1.00-1.23); 1.16 (1.02-1.32); 0.97 (0.94-1.01); 1.47 (1.15-1.89) per SD (33.2 ms); apparent c-statistic 0.61 for SDANN alone, and 0.78 without and 0.80 with SDANN added to the covariates; −6.59 ms (−7.93 to −5.26); 1.16 (1.03-1.29); 1.22 (1.07-1.38); 0.98 (0.94-1.03) |

Abbreviations: AMD, age-related macular degeneration; OR, odds ratio; SDANN, SD of 5-minute average beat intervals.

Halfway to 1 is measured on the log scale from the reference estimate stated in the row, or otherwise from the main estimate, 1.12. Numbers in parentheses are the check numbers in Methods. The 4 later checks (Firth regression to the eye procedure check) were added after the drug-timing analysis, and the final checks (last 5 rows) after all other results had been seen. Readings are shown with the estimates reported in the paper; none differed from the reading first printed.
